# Viral Induced Genetics Revealed by Multi-Dimensional Precision Medicine Transcriptional Workflow

**DOI:** 10.1101/2020.04.01.20050054

**Authors:** Jeremy W Prokop, Ruchir Gupta, Mara L. Leimanis, Derek Nedveck, Rama Shankar, Katie Uhl, Bin Chen, Nicholas L. Hartog, Jason Van Veen, Joshua S. Sisco, Olivia Sirpilla, Todd Lydic, Brian Boville, Angel Hernandez, Chi Braunreiter, ChiuYing Cynthia Kuk, Varinder Singh, Joshua Mills, Marc Wegener, Marie Adams, Mary Rhodes, Andre S Bachmann, Wenjing Pan, Miranda L. Byrne-Steele, D. Casey Smith, Mollye Depinet, Brittany E. Brown, Mary Eisenhower, Jian Han, Marcus Haw, Casey Madura, Dominic J Sanfilippo, Laurie H. Seaver, Caleb Bupp, Surender Rajasekaran

## Abstract

Precision medicine requires the translation of basic biological understanding to medical insights, mainly applied to characterization of each unique patient. In many clinical settings, this requires tools that can be broadly used to identify pathology and risks. Patients often present to the intensive care unit with broad phenotypes, including multiple organ dysfunction syndrome (MODS) resulting from infection, trauma, or other disease processes. Etiology and outcomes are unique to individuals, making it difficult to cohort patients with MODS, but presenting a prime target for testing/developing tools for precision medicine. Using multi-time point whole blood (cellular/acellular) total transcriptomics in 27 patients, we highlight the promise of simultaneously mapping viral/bacterial load, cell composition, tissue damage biomarkers, balance between syndromic biology vs. environmental response, and unique biological insights in each patient using a single platform measurement. Integration of a transcriptome workflow yielded unexpected insights into the complex interplay between host genetics and viral/bacterial specific mechanisms, highlighted by a unique case of virally induced genetics (VIG) within one of these 27 patients. The power of RNAseq to study unique patient biology while investigating environmental contributions can be a critical tool moving forward for translational sciences applied to precision medicine.

**One Sentence Summary:** RNAseq shows the potential of a multidimensional workflow to define molecular signatures for precision/individualized medicine within the pediatric intensive care unit, identifying mechanisms such as viral-induced dominant genetics and infection signatures.

## Introduction

Precision/personalized medicine represents the concept of rapidly identifying altered biology within an individual, using the findings to guide therapy. Much of the promise of this new strategy is obscured by the inadequacy of available technology to discern altered biological pathways in a noninvasive fashion and concomitantly view multiple different facets of altered biology. Whenever a disease process is multi-systemic such as infection, neoplasm, or toxin-mediated, blood has potential to provide early detection to characterize injury. RNAseq has been implemented in many trait mapping projects, using large cohorts of specific phenotypes/trait groups relative to controls to identify biomarkers. It remains to be shown how the millions of dollars of integrated RNAseq work can be translated into individual patients of broad phenotypes to guide precision medicine. The power of RNA generated data likely is in its potential to effect therapeutic changes in individual patients with diverse clinical presentations, such as Multiple Organ Dysfunction Syndrome (MODS). COVID-19 infections highlight this point well, where not every patient with COVID-19 needs to be treated for the MODS that results in lethality, just those that advance into sepsis, where biomarkers of these pathways are critical for earlier treatment of patients.

The Pediatric Intensive Care Unit (PICU) sees diverse phenotypes involving environmental induction (virus, bacteria, trauma), resulting in individual outcomes that could benefit from an expeditious diagnostic and therapeutic timeline. This is ideal for the development and implementation of rapid precision/personalized medicine. Multi-time point analyses that can dissect disease using omic technologies in n=1 biology is currently lacking in the PICU. Patients with MODS have high rates of morbidity and mortality and represent some of the most complex cases within the PICU. Using a combination of PAXgene tubes with ribosomal reduced RNAseq, we present here a dissection of both cellular and acellular RNA signatures of PICU MODS patients, suggesting utility for transcriptomics in precision medicine.

Over the last decade, our ability to extend life in patients with MODS through interventions like dialysis and extracorporeal membrane oxygenation (ECMO) has improved, but there remains an inability to serially track the dyshomeostasis induced by disease from trigger to organ injury, an endpoint for a variety of infectious and noninfectious agents. To halt injury to organs and promote recovery, the clinical team needs rapid, actionable clinical information within the short treatment window of any ICU. That information needs to include simultaneously the etiology, i.e., was it infectious or non-infectious, and the extent of homeostatic disruption. Bacterial cultures take at least 24 hours to grow and 72 hours to speciate. Viral panels only check for common viruses, requiring insights of what to test. Both viral and bacterial analyses are often only ordered when the phenotype is suspected of matching, meaning unique responses are often missed for proper diagnosis. At the same time, the treating physicians are often unsure of the patient’s clinical state, such as whether the patient is immunosuppressed or experiencing an overwhelming immune and inflammatory response *(1–3)*. We are seeing that even in epidemics from an infectious agent like COVID-19, there are differing clinical phenotypes that lead to MODS, requiring changes in therapeutic course *(4, 5)*. In any critically ill patient with MODS, the first 48 hours are spent by the clinical team supporting organ function with little ability to make effective changes to the clinical course. At present, our perception of organ damage is limited to a few organs that have validated, but imperfect means of measurement such as the heart, lung, kidney, liver, and brain, missing markers for many other tissues. The transcriptome is an attractive tool, offering a snapshot of the interaction between affecting pathology and patients’ biology. Thus, transcriptomics is a practical, real-time approach to diagnosis and management, which with decreasing time of processing and improved computational capacity to provide insight, could be a powerful tool to predict disease etiology and clinical course. Doing so reduces diagnostic odyssey in a population that can ill afford delay, providing an agnostic guide to patient-specific disease. The individual responses seen in each MODS patient make this group a prime cohort for personalized/precision medicine strategy development *(6)*, as we lay forth below.

## Results

### Patient Cohort Demographics

The cohort consists of 4 sedation controls (Sedation), 17 MODS, and 6 MODS patients that needed Extracorporeal membrane oxygenation (defined as ECMO throughout). Diverse ethnicities are represented in the data: Caucasian-16, Hispanic/caucasian-6, black-4, east Asian-1, and Hispanic/black-1. While mostly even numbers of male/female in CT and MODS, ECMO patients tend to be male. Renal failure (89%-MODS, 100%-ECMO) and liver failure (30%-MODS 50%-ECMO) are higher in ECMO than MODS group. Eight of the patients were transferred from local hospitals to Helen DeVos Children’s Hospital (HDVCH) to receive more advanced PICU support at a quaternary children’s hospital. A total of 12 patients had been diagnosed with additional comorbidities, including multiple rare diseases diagnosed via genomics. Blood draws were done on each patient followed by RNAseq, with 6 patients having only a day0 transcriptome, 8 with day0/3, and 13 with day0/3/8 transcriptomes (Fig. 1) based on length of stay in the PICU.

**Fig. 1.**
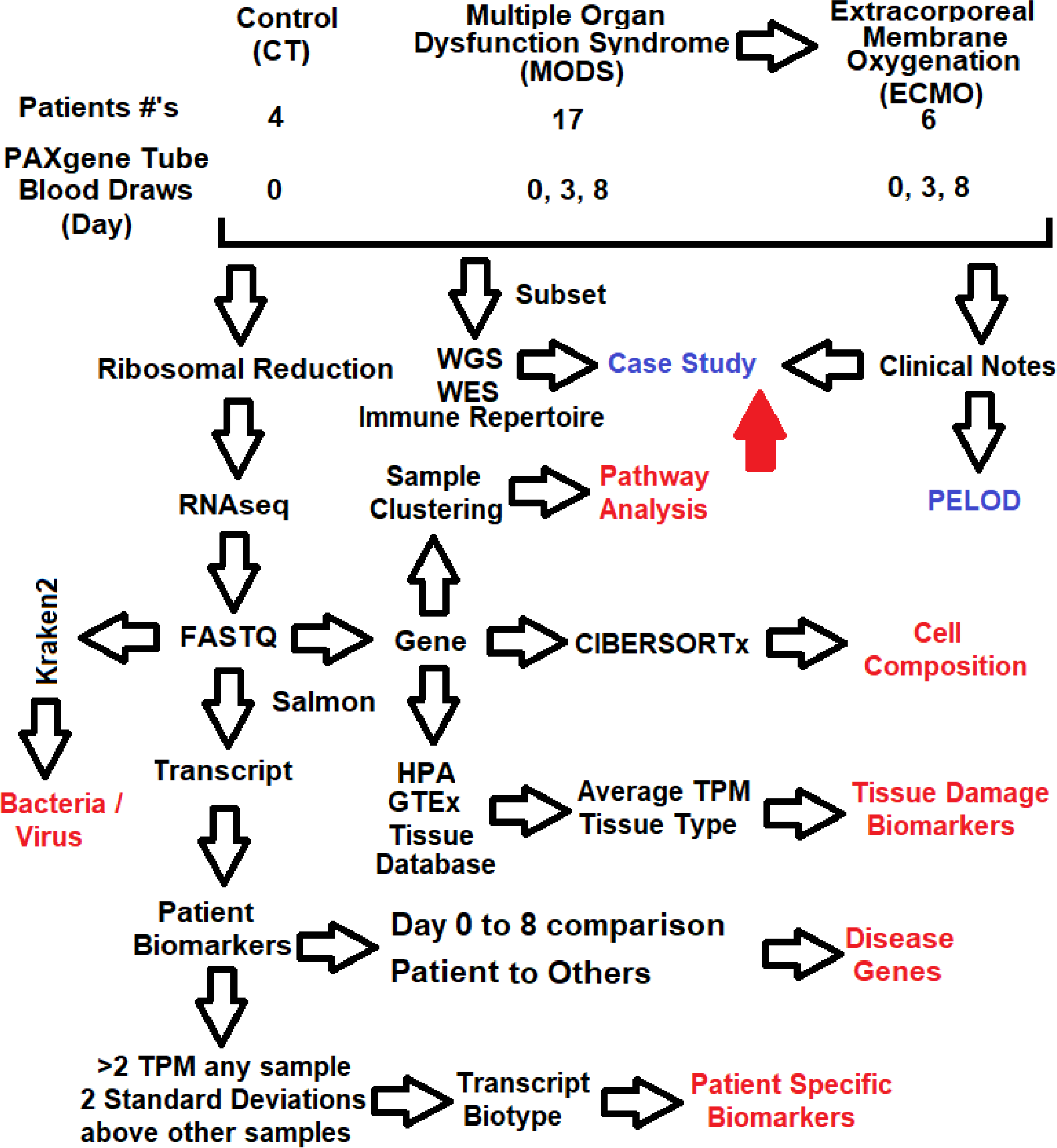
Transcriptomics workflow used to study PICU patient precision medicine. Shown in red are returnable datasets from the RNAseq.

### Transcriptome comparisons to known markers

Going from large cohort studies to individual patients, RNAseq analysis has been very challenging to date. While large sample sizes are used to derive significance and powerful marker genes, it impedes understanding of RNAseq signatures in each patient, mainly when diverse phenotypes are present. The collection of 61 samples from three time points of 27 different PICU patients highlights this. Unsupervised clustering of sample gene-level annotations (27,854) over multiple days shows minimal bulk overlap of the samples (Fig. 2A). Using the day0 RNAseq of the 27 patients, there is some clustering of the sedation controls relative to the MODS/ECMO patients (Fig. 2B).

**Fig. 2.**
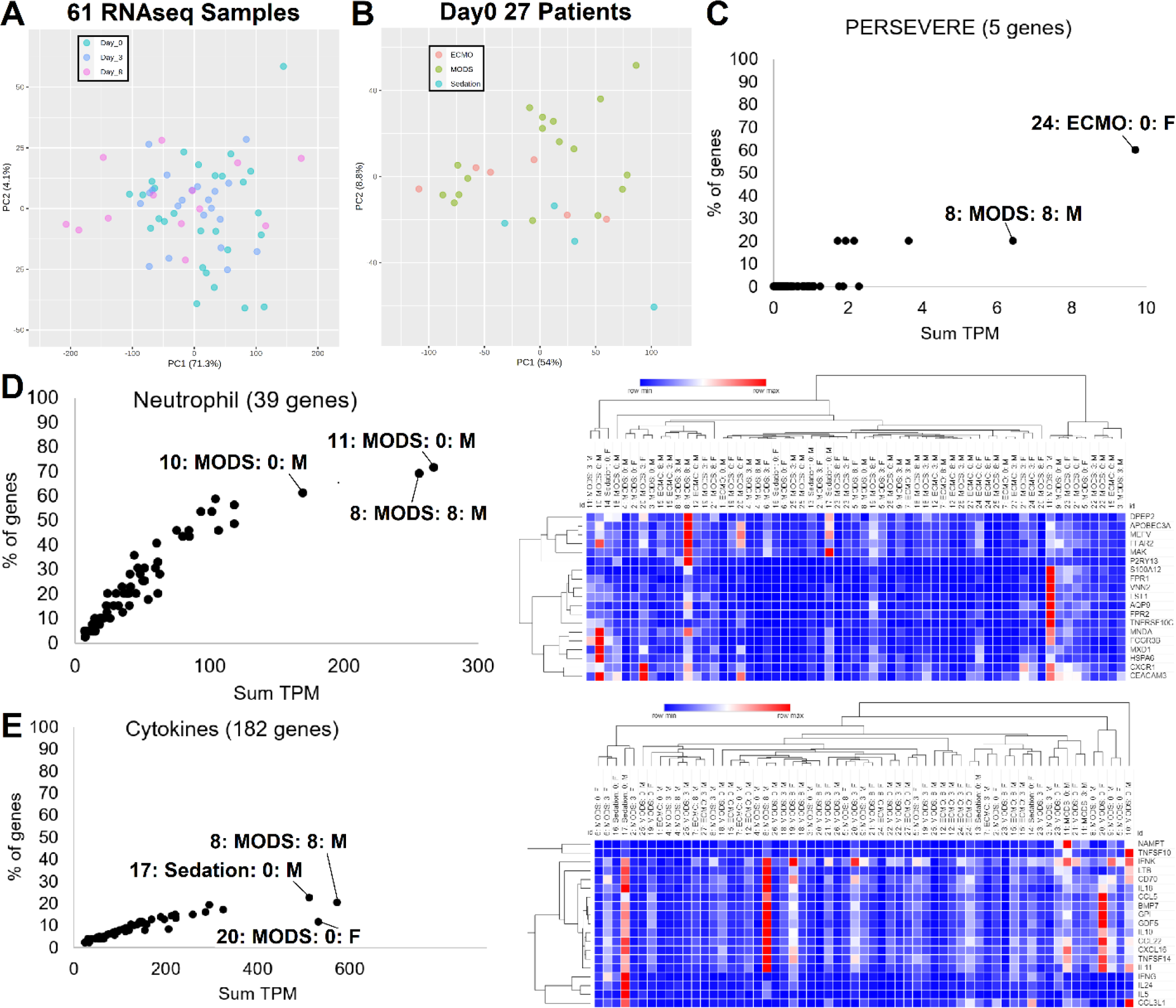
61 transcriptomes analyzed from 27 patients. **(A-B)**. Principle component analysis of transcriptomes annotated at the gene level for all three days (**A**, day0-green, day3-blue, day8-magenta) or on day 0 for the various groups (**B**, Sedation control-blue, MODS-green, MODS with ECMO-red). **(C)**. Using the five PERSERVERE genes, we quantify the total transcripts per million (TPM, x-axis) and the percent of the five genes expressed (y-axis). Two outliers are labeled (sample:group:day:sex). **(D-E)**. Expression of various neutrophil genes (**D**) or Cytokines (**E**) shown on the left as the sum of their TPM (x-axis) and the % of genes in the group expressed (y-axis). On the right is a heat map of each sample and the various genes. Clustering is based on one minus Pearson’s correlation.

Validated biomarkers for pediatric sepsis severity *(7)* were examined and found in two samples to have significant elevation (Fig. 2C), suggesting that sepsis severity is not always the driver of MODS. The two patients with elevated sepsis severity markers had severe outcomes, as seen in patient case descriptions. Another set of genes thought to contribute to MODS are those associated with neutrophil biology *(8)*, where we show the elevation of several neutrophil specific genes *(9)* within our cohort (Fig. 2D). Patient 11 day 0 has the highest neutrophil expression coming from elevation of *S100A12, FPR1, VNN2, LST1, AQP9, FPR2, TNFRSF1, MNDA, FCGR3B* that are connected particularly with neutrophil motility *(10)*. Patient 8 day 8 has the second-highest neutrophil profile based on *DPEP2, APOBEC3A, MEFV, FFAR2, MAK, P2RY13*. Cytokines are known to contribute to MODS *(11)*, where 19 blood cell transcribed cytokines show elevation in patients (Fig. 2E). Patient 8 day 8 has the highest cytokine activation, including *IFNK, LTB, CD70, IL18, CCL5, BMP7, GPI, GDF5, IL10, CCL22, CXCL16, TNFSF14, IL11*. Patient 20 day 0 has the second-highest cytokine activation, including *CCL5, BMP7, GPI, GDF5, IL10, CCL2, CXCL, TNFS, IL11*.

### Cell Composition

From the gene level annotation, we assessed cell composition using CIBERSORTx digital cytometry *(9, 12)*. Of the 22 cell types annotated, there is a broad spectrum of composition predictions (Fig. 3A). Monocytes and neutrophils make up the most substantial portion in most samples, with other cell types having outliers from the cohort averages (Fig. 3B). Two patients on day 3 are outliers for monocyte levels (patients 26 and 27). Neutrophils are highest at day 0 and 3 with much lower levels at day 8, with the inverse seen in CD4 memory T cells resting state. Two patients on day 8 have continued elevated levels of neutrophils (patients 21 and 20). Memory B cells have two samples as outliers on day 0 (patients 21 and 5). Several samples show elevated CD8 T cells, including patient 24 day 0. One patient on day 0 shows elevation of dendritic cells (patient 20) and one on day 8 for Tregs (patient 4). Eosinophil elevation is seen in patient 25 on day 0 and 3, with additional elevation of patient 6 on day 0 and patient 2 on day 3. Four of the patients show elevation of day 0 CD4 memory T-cells, including patient 24 and 19 and two of the sedation controls (17 and 16). Two samples have elevation of mast cells, day 3 patient 24, and day 0 patient 1. Day 0 samples show an elevation of M0 macrophages with highest levels seen in patients 4, 1, and 7 with elevation in patient 22 on day 3 and patients 5 and 15 on day 8.

**Fig. 3.**
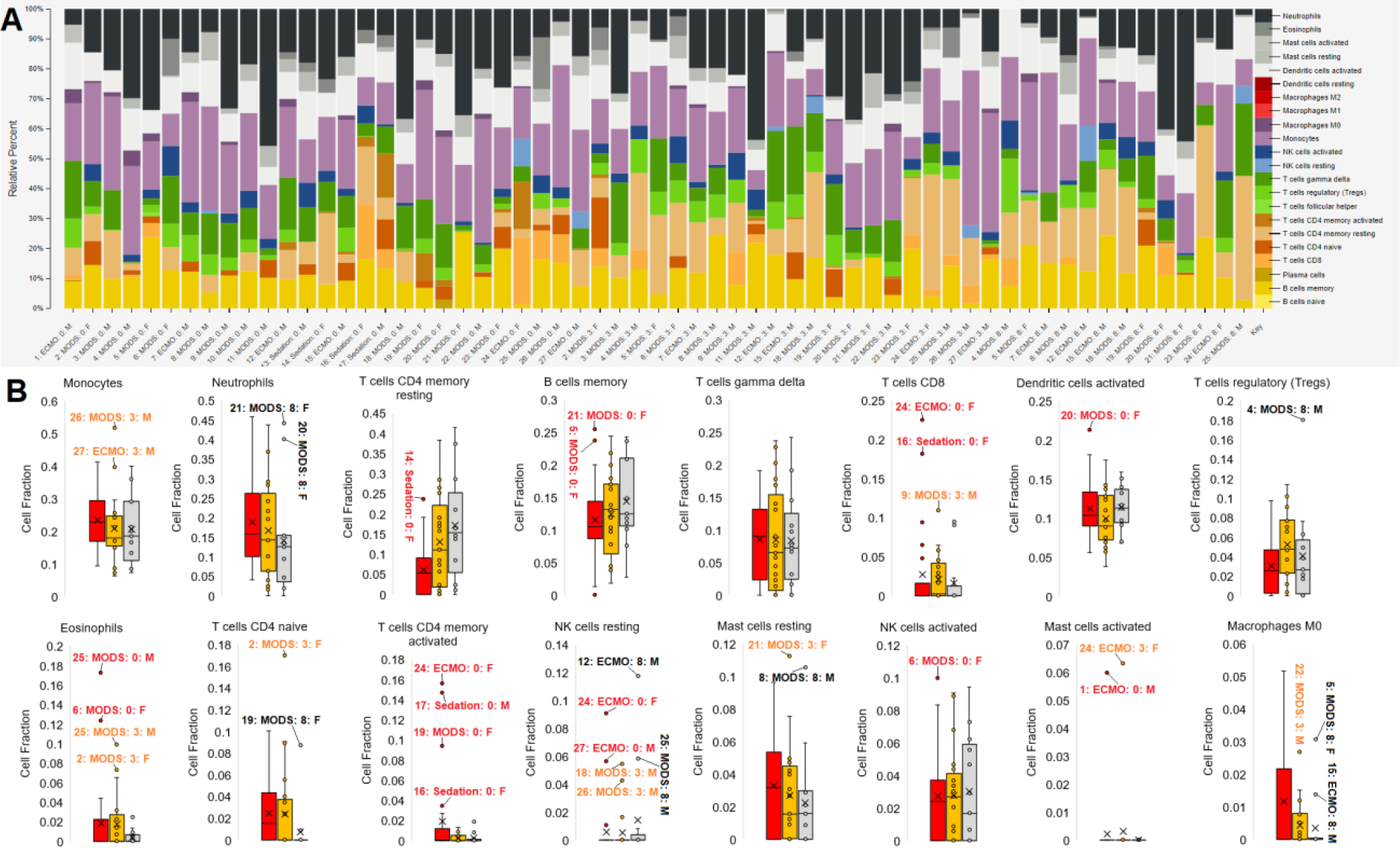
Blood cell composition of each sample. **(A)** CIBERSORTx plot of each sample deconvoluted cell identifies. The color legend is on the far right. **(B)** Break down for 16 of the cell types for day 0 (red), day 3 (orange), and day 8 (gray) shown as box and whisker plots. Outliers are labeled (sample:group:day:sex).

### Bacterial/Viral Mapping

Many of the patients are known to have clinical infections, and the blood draws for RNAseq were performed in PAXgene tubes known to support isolation of bacterial and viral RNA. Therefore, we assessed RNA reads associated with bacteria and viruses using Kraken2, with 20,266 annotations. Mapped reads were normalized to human reads with values around 1,000-2,000 reads for bacteria and viruses (Fig. 4A). Patients 5 and 11 have the highest bacterial read mapping.

**Fig. 4.**
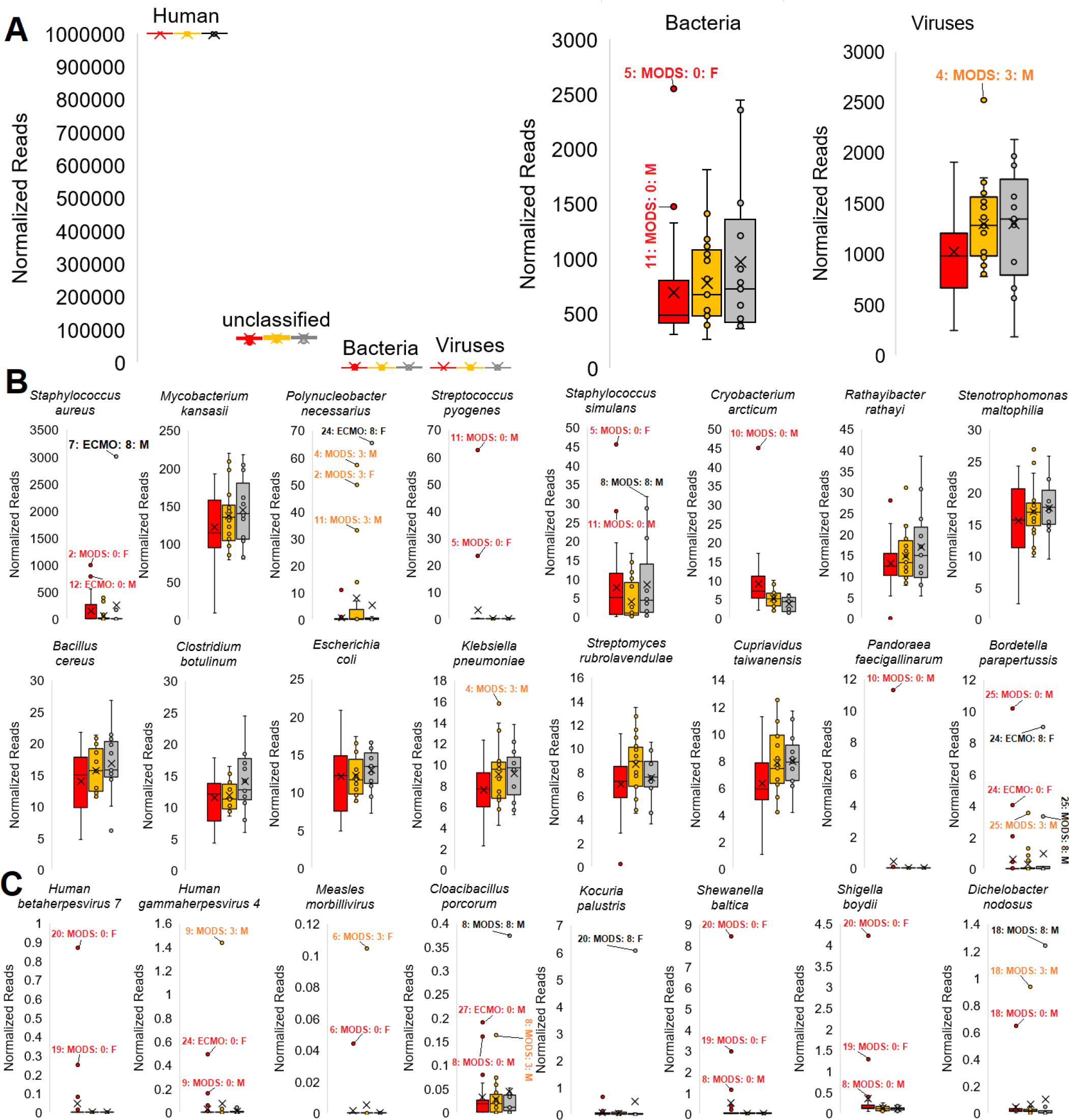
Bacterial/Viral Mapping from RNAseq. **(A)**. Box and whisker plot for domain annotations normalized to every million reads of human for day 0 (red), day 3 (orange), and day 8 (gray). **(B-C)**. Box and whisker plots for the highest mapped bacteria (**B**) and samples with a significant outlier (**C**) reads mapped relative to the number of human mapped reads. Outliers are labeled (sample:group:day:sex).

For bacteria (Fig. 4B), this workflow identified a *Staphylococcus aureus* infection clinically confirmed that showed up at day 8 in patient 7 and day 0 in patients 2 and 12, which are some of the highest values mapped for any species. Several species show up in all samples, including *Mycobacterium kansasii, Staphylococcus simulans, Rathaylbacter rathayi, Stenotrophomona maltophilia, Bacillus cereus*, Clostridium, *Escherichia coli, Klebsiella pneumoniae, Streptomyces rubrolavendulae*, and *Cupriavidus taiwanensis*. Additional insights from the top mapped bacteria include normal flora elevation of *Polynucleobacter necessarius* and *Bordetella parapertussis* in patient 24 suggested to have issues in antigen processing and presentation (case study presented below for *RNASEH2B*), multiple *Streptococcus* strains (including *pyogenes*) identified in patients 11 and 5 that were culture positive, *Pandoraea faecigallinarum* in patient 10, and *Cryobacterium arcticum* in patient 10 day 0.

Focusing on bacteria and viruses that are extreme outliers within one or more samples, we identify *Human gammaherpesvirus 4* (Epstein-Barr, EBV) in patient 24 that was clinically confirmed with viral PCR (Fig. 4C). In certain patients, infections that were unsuspected such as *Measles morbillivirus* (patient 6), *EBV* (patient 9), and *Human betaherpesvirus 7* (patient 20 and 19) that may have been associated with disease outcomes were identified. The *Measles morbillivirus* was the most surprising outcome, with bioinformatics confirming the etiology as that from vaccination and the clinical teams’ investigation of clinical notes revealed a measles vaccine was administered four days before enrollment in the study. Patient 8, a patient with developmental delay and limitation of support, had a suspected infection that clinicians were unable to identify, ultimately resulting in the patient passing away from MODS several days following the study. Using the bacterial mapping, we isolated unique signatures of *Cloacibacillus* in all three days with a further elevation at day 8, a bacteria incredibly challenging to grow and shown to associate with multi-organ pathology similar to this patient *(13)*. While reads align to some of these transcriptomes, reads may belong to closely related species not within our transcriptome database.

### Organ Damage Biomarkers

While assessing the transcripts within the RNAseq samples, we noticed the presence of multiple Albumin (*ALB*) transcripts in patients 24 and 27 at day 0 (Fig. 5A). The *ALB* gene is known to be highly specific to the liver, and its presence was a surprise. Both patients were annotated with liver failure, marked by bilirubin and AST blood elevation. Reads in the *ALB* gene at day 0 of patient 27 cover 89% of the gene transcript with ten or more reads, showing these read annotations are not erroneous (Fig. 5B). Interestingly, both of these patients had a day 3 spike in AST blood levels, not at day 0 when ALB is high (Fig. 5C). This suggests that *ALB* RNA in the blood could have potential to mark liver damage. Moreover, from the human protein atlas, we know that 15 genes are unique to bone marrow. These genes show that all three time points of patient 8 are elevated for blood markers of bone marrow cells, with day 8 the most elevated, and two days elevated in patient 12 (Fig. 5D).

**Fig. 5.**
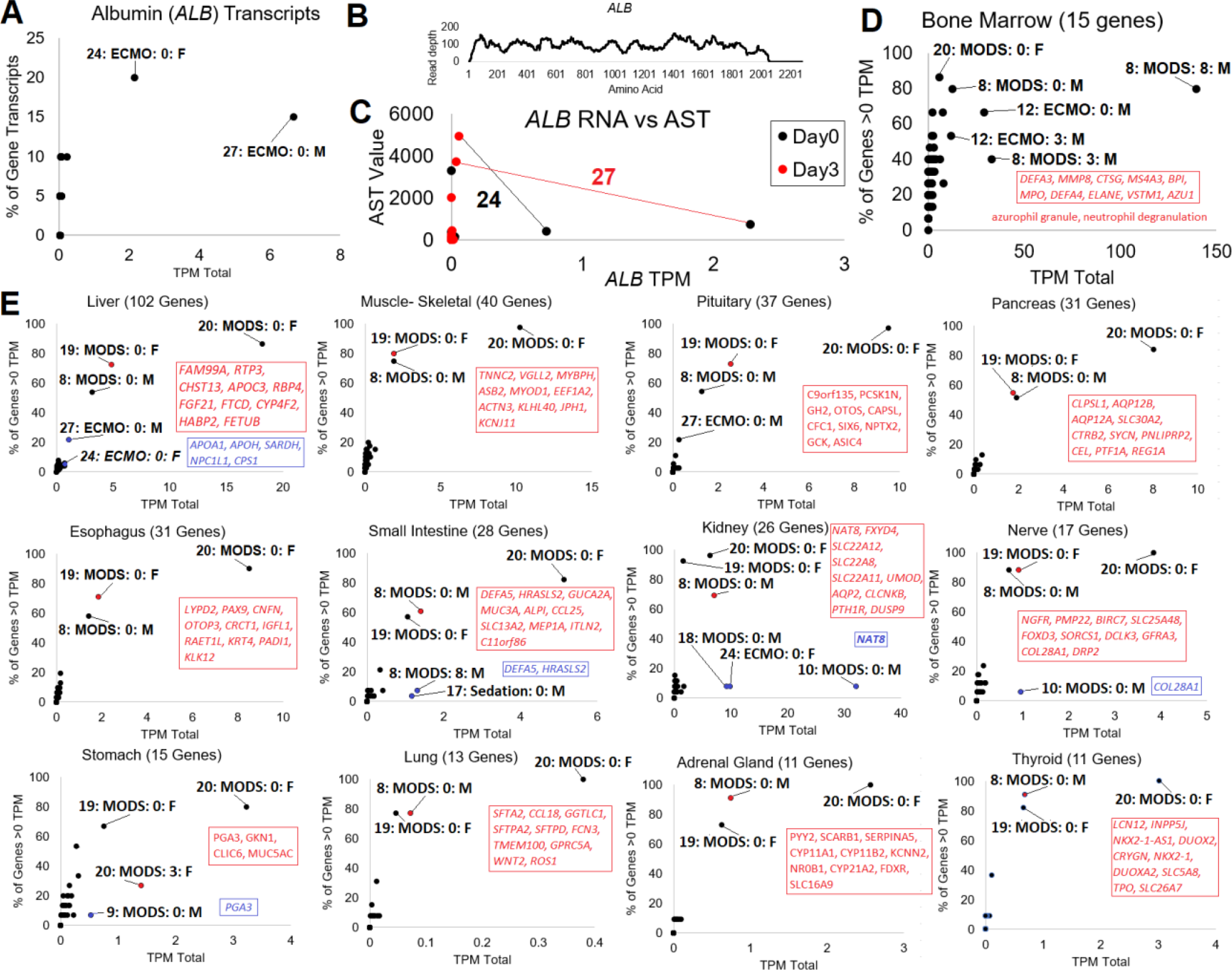
Organ-specific biomarker transcripts. **(A)** Mapping of each RNAseq to transcripts of *ALB*, shown as the summed TPM count (x-axis) and the percent of *ALB* transcripts identified (y-axis). **(B)** BWA alignment of reads from patient 27 day 0 to *ALB* (NM_000477.7). **(C)** Correlation of ALB mapped reads to blood measured AST at day 0 (black) and day 3 (red). The black line connects the two points for patient 24 and red line for patient 27. **(D)** Human Protein Atlas annotated Bone Marrow specific genes assessed in each patient for the summed TPM and the % of genes identified above 0. **(E)** GTEx tissue-specific genes annotated for 12 tissues. Shown for each tissue is the number of genes used for annotations next to the tissue label. The x-axis shows the summed transcript per million (TPM), and the y-axis shows the percent of tissue-specific genes with expression >0. Genes, listed in order of expression value, are shown in red for the top non-patient 20 sample. Genes in blue are shown for small intestine, kidney, nerve, and stomach, where a small number of genes are expressed very high.

To begin understanding the patient-specific tissue responses in each of the transcriptomes, we developed a list of genes that are unique to tissues from GTEx *(14)* and found only a small subset of patient RNAseq to have detectable transcripts for these tissues (Fig. 5E). Three patients are identified as extreme outliers at day0 (patient 20, 8, and 19) in all of the tissues. At day 0, these are the same three patients that show elevation of both *Shewanella baltica* and *Shigella boydii* bacteria in Fig. 4. Top genes identified within these three patients are labeled on the figure. Tissues with additional samples identified include liver, pituitary, small intestine, kidney, nerve, and stomach. Five genes outside of *ALB* are seen elevated in patients 27 and 24 (*APOA1, APOH, SARDH, NPC1L1, CPS1*) for the liver. For small intestine, two genes (*DEFA5, HRASLS2*) are seen elevated at day 8 in patient 8 and day 0 in sedation control patient 17. Three patients at day 0 have elevation of the kidney tubule gene *NAT8* (patient 10, 18, 24). Patient 9, who had necrotizing fasciitis, had the elevation of RNA for *PGA3* that is highly associated with stomach expression.

### Transcript Biotype annotations

A strategy of mapping biological outcomes within each patient was to map transcripts that are two standard deviations above other samples annotated to 226,030 transcripts of Gencode followed by the assessment of transcript biotypes (Fig. 6) and enriched pathways. Again patient 20 day0 RNAseq is an outlier with 18,062 transcripts elevated, followed by patient 6, 8, and 9 as outliers. Annotation of the biotypes identifies several samples enriched, suggestive of altered biology, including patient 20 with retained introns, patient 7 for rRNA, and patient 12 for scaRNA, snoRNA, and mitochondrial tRNA. Utilizing the list of genes elevated for each patient with gene ontology (GO) enrichment suggests multiple overlaps with clinical findings. Enriched genes associated with biological pathways are listed in the case presentations below.

**Fig. 6.**
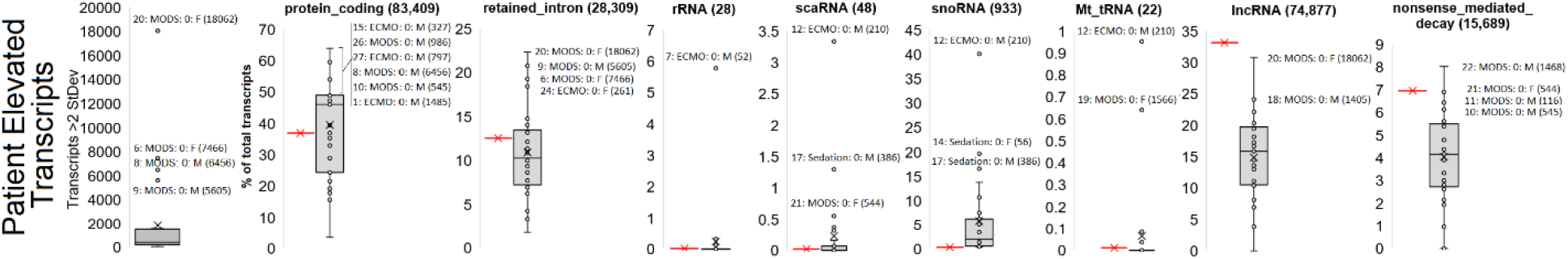
Transcript biomarker outliers. Far-right panel shows the number of transcripts within a transcriptome that are more than two standard deviations from the mean with outlier samples identified. The remaining panels show biotype annotations of elevated transcripts for each patient with the red bar represented the expected frequency of the biotype based on total presence in the transcriptome.

### Building precision medicine dynamic insights from transcriptomics

A total of 13 of the patients had RNAseq for three time points, allowing for comparison of day0 vs. day8 transcriptomes and average of all three time points relative to other patients (Fig. 7). Several of the patients, including patient 20, have an elevation or suppression of many transcripts in day0 relative to day8 (Fig. 7, left) and transcripts elevated in all time points of the patient relative to others (Fig. 7, right). GO enrichment again revealed a remarkable overlap to patient clinical records. Of note, patient 24 had an elevation of 67 transcripts, including multiple miRNA, snoRNA, and snRNA at day0 that were unique to the patient, the only time this occurred, suggesting a unique biological outcome of potential syndromic biology.

**Fig. 7.**
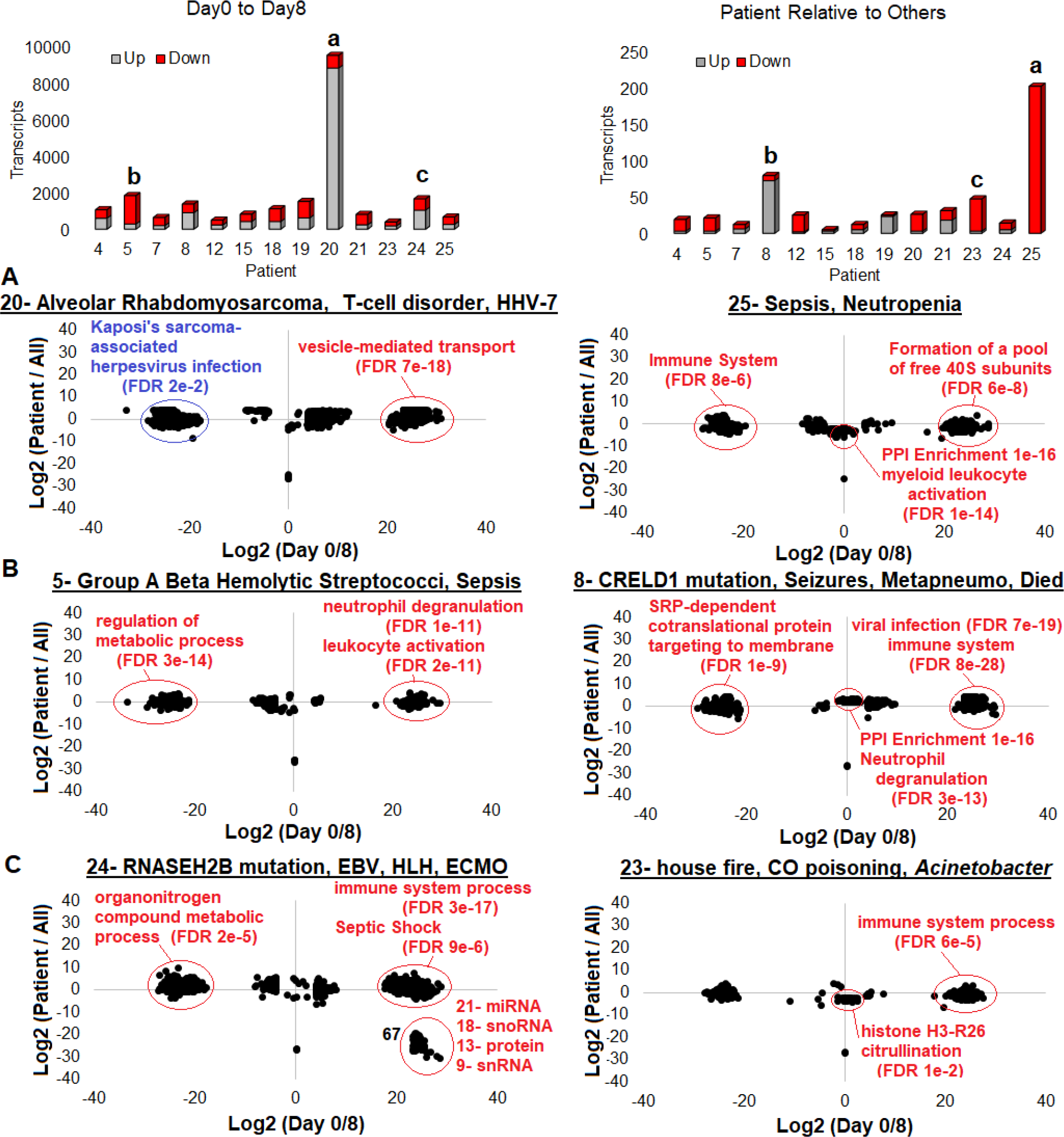
3 Day0-8 and Patient to All genes. **(Left**) Genes up (gray) or down (red) in patients at day0 RNAseq relative to day8. **(Right**) Genes up (gray) or down (red) in all three days RNAseq of patient relative to all other patients. Shown below is the plot of both day0/8 (x-axis) and patient to all (y-axis). Labeled for each patient are the primary diagnoses and the GO enriched terms that overlap to phenotype (red and blue). The most surprising observation was the HHV-7 detected infection in patient 20, had sarcoma, that aligns to genes decreased at day0 that correlate to herpes associated sarcoma.

### Patient precision medicine insights

Using the data for each patient, we detail the 23 MODS/ECMO cases, integrating the transcriptome data with notes and primary diagnosis. Organs in failure are L-liver, R-renal, and C-cardiovascular with MV-mechanical ventilation.

1. (ECMO-Sepsis/Acute myocarditis; R, MV, C; PELOD-12): Clinical Enterovirus and Staph infection; high mapping *Staphylococcus simulants* and *Staph aureus* day0; 1,485 day0 elevated transcripts (GO: ribosomal protein genes*(15)*); elevation of mast cells at day 0; high levels of M0 macrophages at day 0.
2. (MODS-Sepsis/Dilated Cardiomyopathy; L, MV, C; PELOD-11): Clinical Rhino-Enterovirus; high detected *Staph aureus* at day0; 383 day0 elevated transcripts (GO: viral carcinogenesis); elevation of eosinophils and naïve CD4 T cells at day 3.
3. (MODS-Sepsis; L, R, MV; PELOD-12): Clinical OTC deficiency, newborn seizures, no clinically identified pathogens; high day0 mapping of *Pasteurella multocida* and *Tsukamurella paurometabola*; *Pasteurella multocida* has been observed to cause newborn seizures and sepsis *(16)*; *Tsukamurella paurometabola* has been observed to cause sepsis *(17)*; 37 elevated day0 transcripts.
4. (MODS-Sepsis; MV, C; PELOD-11): Died; Sturge Weber syndrome; clinical *Staph* infection; detected *Staph aureus*; enrichment of known chronic based infectious agent *(18) Pandoraea apista* and the highest level of *Klebsiella pneumoniae* at day 3; 601 up (GO: myeloid leukocyte activation) and 465 down (GO: mRNA metabolic process) transcripts day0 to day8; patient elevation of 16 down transcripts in all three days (GO: Defensin/corticostatin family); day 8 spike in Tregs, which are associated with systemic inflammatory response *(19)*; highest level of M0 macrophages at day 0.
5. (MODS-Sepsis; R, MV; PELOD-22): Clinical Group A Beta Hemolytic *Streptococcus*; Enrichment of *Streptococcus pyogenes*; 287 up (GO: neutrophil degranulation) and 1,549 down (GO: metabolic process) transcripts day0 to 8; elevation of memory B cells at day 0; high levels of M0 macrophages at day 8; highest bacterial mapped reads.
6. (MODS-Sepsis; MV; PELOD-11): Schaaf Yang syndrome (MAGEL2 identified variants); clinical Pseudomonas; only patient with detectable *Measles morbillivirus* reads (received vaccine before admission); detection of Pseudomonas phage PaP1 and *Pseudomonas entomophila* a known hemolytic soil bacterium *(20)*; 7,466 day0 elevated transcripts (GO: ribosomal protein genes*(15)*); elevation of eosinophils at day 0.
7. (ECMO-Sepsis; R, MV; PELOD-20): Vascular Sling; Clinical *Staph* infection; Day8 highest *Staph aureus* detection; 199 up (GO: Immune System) and 443 down (GO: cellular process) transcripts at day0 to 8; high level of M0 macrophages at day 0.
8. (MODS-Sepsis; MV; PELOD-20): Died; Seizure disorder with identified variants in *CRELD1* (p.Q320RfsX25, p.C192Y); Clinically confirmed Metapneumo; *Raoultella* and *Cloacibacillus* enriched in all three days of RNAseq with *Cloacibacillus* highest at day8 (*Cloacibacillus has been associated with multi-organ pathology in multiple cases (13)*); elevation of 72 transcripts at all days relative to other patients suggestive of neutrophil degranulation (1e-12), leukocyte activation (FDR 6e-13), and linked with known neutrophil protease associated organ pathologies *(21–24)*. Second highest PERSEVERE gene elevation. Neutrophil activation of *DPEP2, APOBEC3A, MEFV, FFAR2, MAK, P2RY13*. Cytokine activation of *IFNK, LTB, CD70, IL18, CCL5, BMP7, GPI, GDF5, IL10, CCL22, CXCL16, TNFSF14, IL11;* day 0 elevation of both *Shewanella baltica* and *Shigella boydii;* spike in bone marrow specific genes at all three days with the highest at day 8; elevation of tissue damage biomarkers at day 0.
9. (MODS-Sepsis; MV; PELOD-12): Necrotizing fasciitis; clinical *Streptococcus pyogenes* infection, aspartate aminotransferase: 45 units/L, albumin: 2 g/dL indicating possible liver damage; high mapping of *Clostridum perfringens* on day0 and 3, *C. perfringens* is associated with necrotizing fasciitis and hypoxia *(25)*; elevation of *PGA3* associated with stomach at day 0; 5605 elevated transcripts of which some indicate HIF-1 signaling pathway enrichment involved in metabolic adaptation to hypoxia; abnormally large proportion of elevated transcripts are retained introns; spike in mapping of human gammaherpesvirus 4 on day 3 and observed at day 0 at lower levels.
10. (MODS-Sepsis; MV; PELOD-5): Clinically identified coronavirus; day0 high mapping of *Pandoraea faecigallinarum* known to be a chronic lung colonizer *(26)* and several *Staphylococcus* infections; high level of mapped reads to *Cryobacterium arcticum* at day 0; high level of *NAT8* at day 0 associated with kidney tubule cells; spike in *COL28A1* at day0 associated with nerve tissue.
11. (MODS-Sepsis; R, MV; PELOD-11): Clinical *Streptococcus pyogenes* infection; day0 high mapping of *Streptococcus pyogenes* as well as other *Streptococcus* infections; day0 high mapping of *Listeria monocytogenes* and *Staphylococcus aureus;* Neutrophil activation of *S100A12, FPR1, VNN2, LST1, AQP9, FPR2, TNFRSF1, MNDA, FCGR3B;* second-highest day 0 bacterial mapping.
12. (ECMO-Sepsis; R, MV; PELOD-11): Clinical human orthopneumovirus; day0 high mapping of *Staphylococcus aureus* and *Burkholderia*; spike in bone marrow specific genes at days 0 and 3; 23 decreased transcripts compared to other patients (GO: *Staphylococcus aureus* infection); 210 elevated day0 transcripts of which snoRNA, scaRNA, and mt tRNA are abnormally high.
13. (ECMO-Sepsis; L, R, MV; PELOD-11): Clinical haemophilus infection and human orthopneumovirus infection; day0 high mapping of *Haemophilus influenzae, Acinetobacter baylyi* (known to be opportunistic with other pathogens *(27)*), and *Pasteurella multocida* (known infection from animal scratches*(28)*); high level of M0 macrophages at day 8.
14. (MODS-Sepsis; R, MV; PELOD-20): Clinical *Serratia mercescens*, rhino/enterovirus, coronavirus; all three day high mapping of *Dichelobacter nodosus*; day0 high mapping of *Paenibacillus* sp. FSL R5-0912 and *Lysinibacillus fusiformis*, which have been observed to lead to sepsis *(29)*; high mapping of *Staphylococcus aureus;* high level of *NAT8* at day 0 associated with kidney tubule cells; 419 transcripts increased from day0-8 (GO: immune system process).
15. (MODS-Sepsis; R, MV, C; PELOD-11): Clinical MRSA, *Staphylococcus aureus*, rhino/enterovirus infection; day0 high mapping of *Staphylococcus simulans* and human betaherpesvirus 7; day 0 elevation of both *Shewanella baltica* and *Shigella boydii;* elevation of tissue damage biomarkers at day 0.
16. (MODS-Acute Kidney Injury; R, MV; PELOD-11): Clinical Alveolar rhabdomyosarcoma and common variable immunodeficiency with predominant immunoregulatory T-cell disorders; high mapping human betaherpesvirus 7 (HHV-7) day0 known to infect CD4+ T-cells *(30)*; day 8 elevation of *Kocuria palustris* that is known to be associated with catheter-related bacteraemia *(31)*; 18,062 day0 elevated transcripts, high proportion of which are retained introns suggesting cancer *(32)*; 700 transcripts down day0-8 (GO: Kaposi’s sarcoma-associated herpesvirus infection); HHV-7 z-score day0/3/8: 6.2/-.2/-.2; Cytokine activation of *CCL5, BMP7, GPI, GDF5, IL10, CCL2, CXCL, TNFS, IL11*. Day 8 continued elevation of neutrophils; elevation of dendritic cells at day 0; day 0 elevation of both *Shewanella baltica* and *Shigella boydii;* elevation of tissue damage biomarkers at day 0.
17. (MODS-Sepsis; MV; PELOD-11): *MECP2* duplication at Xq28 region; clinical Rhino-Enterovirus; clinical aspartate aminotransferase: 56 units/L, albumin: 2 g/dL indicating possible liver damage; *MECP2* duplication has been linked to immunodeficiencies *(33)*; Day 8 continued elevation of neutrophils; elevation of memory B cells at day 0.
18. (MODS-Sepsis; MV; PELOD-11): Clinical Staph Infection; 25% burns that developed sepsis; Clinical aspartate aminotransferase: 58 units/L, albumin: 1.1 g/dL; WBC: 38*10^9^/L; *Halothece* mapped Cyanobacteria that can have cyanotoxins that yield similar outcomes observed in patient namely hepatocyte necrosis *(34, 35)*; known 1468 day0 elevated transcripts (GO renal allograft rejection*(36)*); high level of M0 macrophages at day 3.
19. (MODS-CO toxicity; MV; PELOD-10): Clinical Rhino-Enterovirus; high mapping of *Acinetobacter* and *Burkholderia* infections on day0 and 3; 44 transcripts low at day0 (GO: histone H3-R26 citrullination). H3-R26 citrullination is involved in increasing immune response *(37)*; 180 transcripts increased from day0-8 (GO: immune system process).
20. (ECMO-HLH/EBV; L, R, MV; PELOD-10): hemophagocytic lymphohistiocytosis, EBV detected by PCR and seen in RNAseq of patient at day0; elevation of Bordetella parapertussis at day 0 and 8; Enriched for genes involved in T-cell activation; Highest PERSEVERE gene elevation at time point 0 in cohort; Highest level of CD8 T cells and memory CD4 T cells; elevation of mast cell genes at day 3; elevation of blood *ALB* transcripts at day 0 with five additional liver genes; high level of *NAT8* at day 0 associated with kidney tubule cells; Patient case write up described below.
21. (MODS-Sepsis; L, R, MV; PELOD-22): Clinical hemophagocytosis in bone marrow, clinical Rhino-Enterovirus; high mapping of *Methylobacterium (38), Streptomyces (39)* on day0 and 3; high mapping of *Bordetella parapertussis (40)* on day0; 201 transcripts low on day0 (GO: myeloid leukocyte activation); 265 transcripts increased from day0-8 (GO: translation, cotranslational protein targeting to membrane, GTP hydrolysis and joining of the 60S ribosomal subunit); elevation of eosinophils at day 0 and 3;
22. (MODS-Febrile infection-related epilepsy; L, MV; PELOD-32): *POLQ* and *SASH3* variants; clinical febrile infection-related epilepsy; liver failure; high mapping of *Mycobacterium tuberculosis* and *Pasteurella multocida*, both pathogens have been observed to cause seizures *(41, 42)*; Day3 elevated monocytes.
23. (ECMO-Sepsis/Myocarditis; L, R, MV, C; PELOD-12): Died; clinical myocarditis and culture-negative endocarditis; clinical *Staph* infection; liver failure; renal failure; brain injury; respiratory failure; high mapping of *Pasteurella multocida* (common infection from cat scratches) and *Gordonia polyisoprenivorans VH2, Pasteurella multocida* has been observed to cause endocarditis as well as respiratory tract symptoms*(43, 44)* and *Gordonia polyisoprenivorans VH2* has been observed to cause endocarditis *(45)*, family had two cats as pets; Day3 elevated monocytes; *ALB* RNA detected in blood at day 0 with five additional liver genes; several pituitary associated genes elevated.

### A case of viral-induced genetics (VIG), patient 24

Patient 24 was suggested to have unique potentially syndromic biology based on data presented, agreeing with clinicians who ordered rapid whole-exome sequencing. A previously healthy, active 16-year old female was admitted with fever, malaise weakness, and upper respiratory symptoms that progressed rapidly into multi-organ failure (Fig. 8A). The symptoms started about ten days before hospital presentation and were mainly respiratory. She was initially treated at home with a 10-day course of Amoxicillin. Over the next week, she continued to deteriorate, with on and off febrile episodes thought to be “flu-like” with significant myalgia, eventually becoming bedridden with altered mental status leading to an ED visit. She continued to deteriorate despite cardiac support and required ECMO within 24 hours of being admitted to HDVCH PICU. The rapid deterioration was suggestive of viral illness (identifying EBV by PCR), a ferritin level >10,000 ng/L, and a bone marrow exam suggested Hemophagocytic Lymphohistiocytosis (HLH), a diagnosis that typically takes several days to make but is visible in day0 RNAseq. The patient samples cluster relative to CT and a two year follow up transcriptome (Fig. 8B). A second tool confirmed EBV reads from RNAseq, including the gene *EBER1* (Fig. 8C). Rapid whole-exome sequencing identified compound heterozygous mutations in *RNASEH2B* that GeneDx annotated as likely pathogenic for Aicardi-Goutières, V20L (paternal), and a potential splicing variant c.245-8A>G (material). Aicardi-Goutières is typically an early-onset autoinflammatory systemic lupus erythematosus *(46)*, which is commonly associated with HLH. The V20L variant based on our computational tools has no alteration of the protein (Fig. 8D-E), suggesting it to be benign, and therefore, the patient does not have homozygous associated Aicardi-Goutières. However, the V20L variant was used to show an allelic imbalance from the RNAseq that was pronounced on all but day0 (Fig. 8F), suggesting that a unique form of Aicardi-Goutières may be present.

**Fig. 8.**
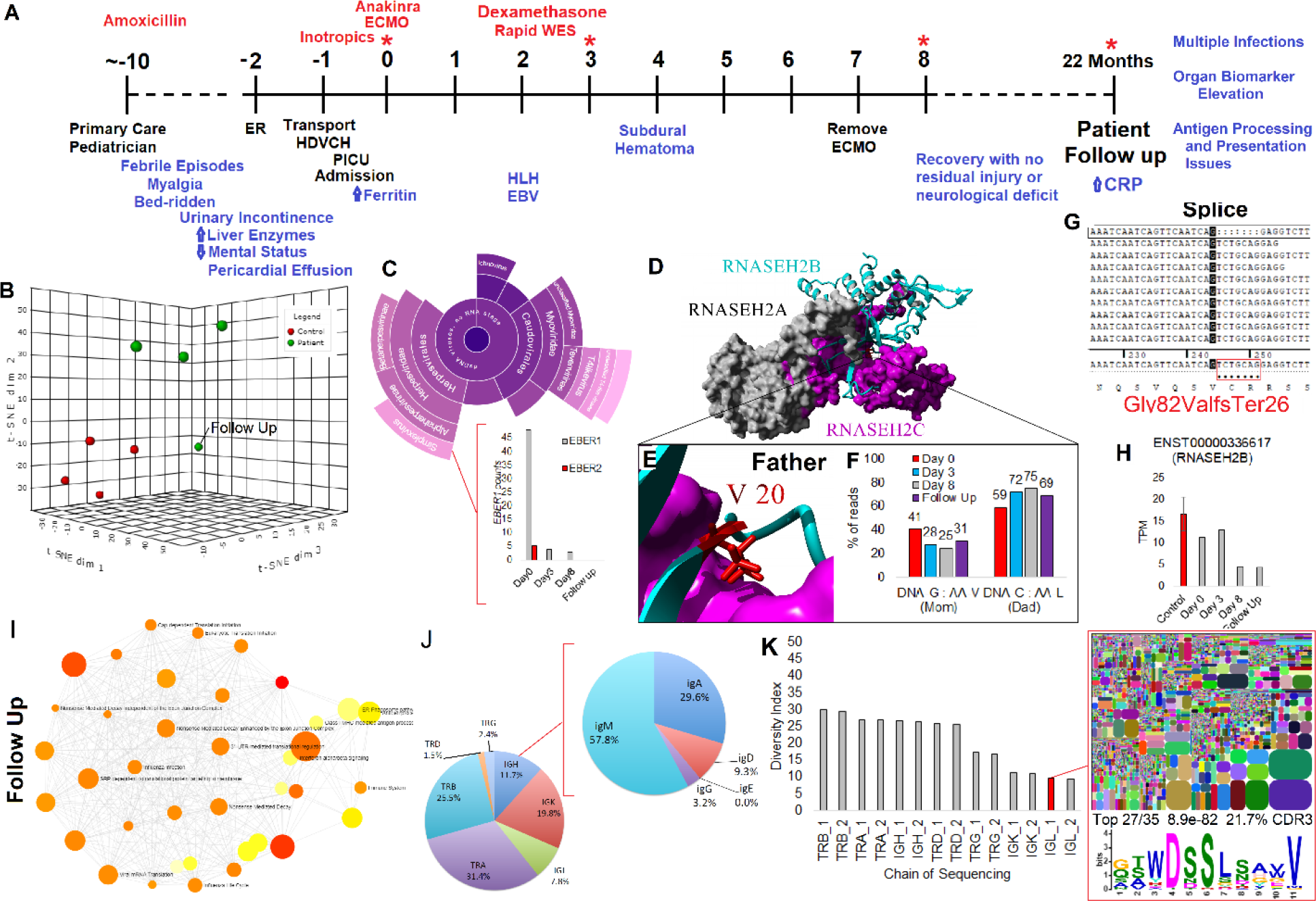
Patient with active EBV and RNASEH2B splice variant: **(A**) Patient timeline: * marks multi-omic data collection points, red marks interventions, black marks major events, and blue marks phenotypes observed. **(B**) Clustering of control (red) and patient samples (green), including a 22 month follow up. **(C)** Second viral read caller to confirm EBV infection with EBER1 (gray) and EBER2 (red) reads from EBV detected at day0 shown below. **(D)** Known structure of RNASEH2A (gray), RNASEH2C (magenta), and RNASEH2B (cyan). **(E)** Zoom-in view of V20 not falling at a critical site suggesting L would be functional. **(F)** Percent of reads from the RNASEH2B alleles at multiple RNAseq measurements suggesting allelic bias to the allele inherited from the father. **(G)** Reads aligned near the potential splice site variant inherited from mom that confirms a frameshift and early truncation. **(H)** Total RNASEH2B levels over the multiple days suggesting reduced levels relative to controls (red). **(I)** Pathways activated in the patient’s two year follow up, suggesting immune system dysfunction. **(J)** Immune system repertoire analysis performed on the patient follow up for seven chains (TRB-T-cell receptor beta; TRA-T-cell receptor alpha; IGH-Immunoglobulin heavy chain; TRD-T-cell receptor delta; TRG-T-cell receptor gamma; IGK-Immunoglobulin light chain kappa; IGL-Immunoglobulin light chain lambda). Shown is the percent of CDR3 identified in each group with a call out for further dissection of the IGH subgroups. **(K)** Diversity index (DI) for each of the sequenced chains. DI corresponds to elevated recombination sites, with lower levels suggesting an enrichment of certain sequences within the chain. The IGL is shown for diversity, suggesting the elevation of epitope recognition. This epitope is seen in 27 of the top 35 CDR3 sequences for IGL with a significant enrichment (8.9e-82) and composing 21.7% of all the CDR3 IGL sequences.

Interestingly, the splice variant inherited maternally (c. 245-8A>G) was expressed only in the day0 samples, yielding a frame-shift mutation (Fig. 8G). Levels of *RNASEH2B* were around 50% in later days (Fig. 8H), suggesting the splice variant had a dominant-negative effect, which would be normally suppressed by nonsense-mediated decay (NMD). This loss of NMD in day0 sample is interesting and correlates with the increasing copies of serum EBV noted clinically and in the RNAseq. Interestingly, it is known that EBV coded proteins can suppress nonsense-mediated decay *(47)*. With the patient’s potential syndromic biology identified, the patient had a two year follow up transcriptome analysis. This revealed excessive inflammation and continued alterations in the immune system (Fig. 8I). The immune system was profiled by sequencing the CDR3 region of seven chains, identifying a high level of IgD (Fig. 8J), and low diversity of immunoglobulin kappa and lambda chains with enrichment of a functional motif (Fig. 8K). Hyper-IgD syndrome can be caused by mutations in MVK, and this gene was reanalyzed in the exome and showed 100% coverage at a minimum of 10X with no variants identified. This all suggests that the patient has extensive immune system dysfunction that, combined with an active EBV infection turning on the suppressed *RNASEH2B* splice variant to cause a dominant-negative HLH that recovered as the EBV was cleared. This patient represents an example of the complex interplay between environment and genetics that we coin as viral induced dominance and highlights the power and need for precision transcriptomic analysis performed on more patients, with these cases missed in large cohort-style study design.

## Discussion

Transcriptomics has been speculated to have significant utility for personalized/precision medicine *(48, 49)*. While fields like oncology have embraced these tools, their value remains undetermined for most other areas in health care. Patients admitted to the intensive care units have altered homeostatic pathways that result from unclear triggers. Transcriptomics has the potential not only to identify the degree of disruption but identify etiology in patients that often don’t have the luxury of time for a long diagnostic odyssey. Thus far for PICU patients, strategies have focused on developing larger cohorts based on similarity of immune responses, typically utilizing microarray strategies focusing on those that have sepsis or MODS *(50, 51)*. In these models of research, clusters of genes are examined for similarity of response and highlighted, where the differences between individual patients are either ignored or not studied.

In the clinical arena, multiple aspects of disease etiology and outcomes exist, and as the technologies for understanding sepsis and MODS biomarkers has evolved, there is much space for developing tools that have potential to actually impact therapy *(52)*. A typical cohort of PICU patients presenting MODS, as shown in this paper, is composed of broad etiology of disease, including diverse environmental contributions and various infections. As the case reviews of these patients show, multiple factors are also identified in many of these patients, from genetics and infection to multiple infections sometimes in each patient. Even within an extensive cohort collection, the combinations that give rise to pathologies in each patient are likely such that no two patients present the same. In this work, we show how the collection of blood samples into the PAXgene tube system, followed by reduced ribosomal RNAseq can elucidate MODS understanding using outlier mapping strategies. The information collected from transcriptomics reveals a remarkable ability to map pathologies and understand the diverse biological responses seen within each patient in the PICU, which gains more insights as the cohorts of information grow. In the age of expanding sequencing technologies and precision medicine, this work highlights that it is important to consider how each patient is unique and to not lose this data by overly investing into cohort based patient assessments that tend to blind individual unique outcomes and mechanisms of pathology. This is further exemplified by viral outbreaks, such as COVID-19, showing selectivity in age and sex related vulnerability, inflammatory phenotypes leading to differences in presentation, severity, and management *(4, 53)*. These differences may result from common factors or could be contributed by unique biology that is challenging to capture in large cohorts, especially if there is no in-depth analysis in a patient-by-patient manner.

One of the most promising parts of the PAXgene collection and its analysis could be in pathogen detection, as the sequencing uncovered microbial/viral RNA that were in the sample and correlate well to clinical confirmed pathogens. This emerging field, labelled metagenomics, is still in the fledgling phase as we are trying to understand its clinical implications *(54)*. The potential for clinical relevance is undeniable, as we showed clinical overlap in multiple patients not only detecting viruses such as measles and EBV, but also bacteria confirmed by clinical tests and those that cannot be cultured well in clinical labs. The power of such findings is in the potential to guide clinical decisions on day 1 as shown by others through discovering unexpected infections *(55)* and antimicrobial choice *(54)*.

As we sequence more patients, we will have more cases that align phenotype with correlating transcriptomic insights. This will require blood draws from larger numbers of patients collected using similar protocols, but with a focus on understanding each patient instead of the entire cohort. Investments into larger datasets should lead to the scaling of bioinformatic tools that will delineate initiation events, patient responses, and determine when and why patients respond adversely. One limiting factor that needs to be surmounted is the current bottleneck by the time it takes to generate RNAseq data using existing platforms. Newer tools such as Nanopore hold the promise for speed to clinical care to transition from observational science, as in this study, into a tool that generates personalized/precision data in real-time, leading to the appropriate confirmatory clinical tests to guide decision making in the PICU.

## Methods

### Sample Collection

IRB approval (2016-062-SH/HDVCH) was obtained for precision medicine initiative to characterize patients admitted to our PICU with multi-organ dysfunction. Blood samples were collected at up to 3 independent time points, at baseline (day 0), greater than 48 hours to less than 72 hours (day 3), and greater than 7 days (day 8). These time points were collected to reflect the trajectory of critical illness with measurements in the acute, stabilization, and recovery phase of illness. The clinical team extracted from the clinical notes, diagnosis, transfer status from other hospital, genomic diagnosis/comorbidity for patient (if available), two-year history within the hospital system, PELOD score, race, age, sex, clinically identified infections, organs experiencing failure, and several clinical measures when available.

### Patients

The sedation patients were less than 18 years of age presenting for routine sedation in an outpatient clinic without an inflammatory disease. The definition for MODS was taken from the original definition *(56)* in which among other criteria, MODS patients were clinically identified with two or more organs in failure. ECMO was MODS patients that required ECMO therapy.

### PAXgene RNA

PaxGene® RNA tubes were stored at room temperature for at least 2 hours following the addition of blood, overnight at -20 °C, and at -80 °C for long-term storage. PAXgene tubes were thawed at room temperature for 2 hours and processed using Qiagen’s QIAsymphony PAXgene Blood RNA Kit.

### RNAseq

RNAseq libraries were generated using RiboZero-Globin rRNA kit (Illumina), KAPA RNA HyperPrep Kit (v1.16) (Roche Life Sciences), and Bio Scientific NEXTflex Adapters (Bioo Scientific). Libraries were pooled and run over 7 flowcells using 150 bp sequencing kit (v2) in paired-end on an Illumina NextSeq 500.

### Human Bioinformatics

Paired-end reads (fastq) were aligned in salmon_0.14.1 *(57)* using the *Homo sapiens* Gencode 31 for transcripts or our custom single gene to sequence transcript library for genes. Mapped transcript per million (TPM) for all samples were processed through NetworkAnalyst3.0 *(58)* using DESeq2 *(59)* to identify gene-level annotations that differ between control, MODS, and ECMO groups. Pathways identified in Network Analyst to have variable gene expressions with an adjusted p-value (< 0.05) were extracted for Gene Ontology and KEGG associated genes. Pooling all day0 TPM values for transcripts, z-scores were calculated to identify day0 samples with greater than 2 standard deviations above the mean of all and a value >1 TPM. The biotype data for the transcripts in each of these samples >2 standard deviations were compared to the % of biotype annotations in the entire transcriptome to calculate transcript enrichment. The genes corresponding to the transcripts >2 standard deviations were assessed with STRING *(60)* for pathway enrichment. For patients with transcriptomes at day0 and day8, Log2 fold change was calculated for day0 relative to day8 on that patient and compared to day0 of the patient relative to day0 of all other patients. Neutrophil specific genes were taken from the LM22 matrix of CIBERSORTx *(9, 12)* and cytokines identified from protein annotations on UniProt for cytokines. Heatmaps were created using the Broads Morpheus tool with each gene normalized over all samples, and clustering performed using one minus Pearson correlation. CIBERSORTx digital cytometry cell fraction imputation *(9, 12)* was performed using the gene mapping on the LM22 signature matrix without batch corrections and 500 permutations.

### Infection Bioinformatics

Using only the first of the paired-end reads, bacterial and viral read mapping was performed using kraken2-2.0.7-beta *(61)* against the full transcriptome database of the tool downloaded on June 24, 2019. Samples were normalized to every million mapped reads to human and clustered based on patient ID. Z-scores were calculated for each viral or bacterial species annotations across all time points for all patients to determine outliers. When needing a secondary read alignment, BWA *(62)* alignment to individually downloaded transcriptomes were obtained.

### Organ Biomarker database

Using GTEx *(63)* tissue annotated expression datasets, we filtered for genes that are unique to each tissue/organ. All sex-specific organs were removed from the database to begin. Genes had to have an expression value >5 and an enrichment of 10-fold in one tissue over the average of all. Each annotated tissue needed to have at least 5 genes identified to be included in the search. From the patient samples included genes for annotation had to have a 20 fold elevation in one of the samples relative to the average of all samples. The TPM values for each tissues were added together for sample and the percent of genes with a value above 0 determined.

### RNASEH2B patient assessment

A detailed analysis of patient 24 was performed. A 2 year follow up PAXgene tube sample was isolated from the patient and RNA extracted as before. Illumina RNAseq was performed on the isolated RNA as before. Leftover RNA was run on Nanopore RNAseq using PCR-cDNA Sequencing kit and MinION flow cell, comparing values to RNA obtained from a control sample. Illumina and Nanopore differentially expressed transcripts were compared to isolate overlapping data, running the gene list through NetworkAnalyst3.0 for enriched pathways. The last bit of isolated RNA was sent to iRep for Immune Repertoire analysis of seven chains of CDR3 of the immune system. Reads from day0 were reassessed for viral mapping using Taxonomer (www.taxonomer.com) and the EBV transcriptome aligned with BWA*(62)*. Rapid whole-exome sequencing on the patient, and both parents was performed clinically through GeneDx. Assessment of variant outcomes in RNASEH2B was done using our previously published workflow*(64)*. Reads from all of the patient’s RNAseq were aligned onto the RNASEH2B transcript using BWA*(62)* followed by calculation of allelic bias using the variant inherited from the father. Reads were then manually viewed in Sequencer software to interpret the splice variant site for protein prediction.

## Data Availability

Material for gene, transcript, and Kraken mapping can be found at https://drive.google.com/file/d/1Tr0zEdAmRY2CdcTn6-MX62XC2MpSeXDt/view?usp=sharing

## Acknowledgments

We want to thank the patients for their samples, and the PICU staff at Helen DeVos Children’s Hospital for their support with the sample collections and their various contributions.

## Author contributions

Performed sample prep and sequencing (KU, TL, MW, MA, MR, WP, MLBS, DCS, MD, BEB, ME), analyzed data (JWP, RG, DN, RS, JVV, JSS, OS, CK, VS, JM, SR), organized clinical information (MLL, NLH, BB, AH, CB, MH, CM, DJS, LHS, CB, SR), wrote manuscript (JWP, RG, MLL, SR), oversaw project completion (JWP, BC, ASB, JH, DJS, SR), all authors have approved of the manuscript submission.

## Data and materials availability

All of the mapping data for the RNAseq are available for gene and transcript levels in the supplemental Excel file.

